# Older drivers, family members and caregivers’ views on the NSW age-based licensing; a mixed-methods approach

**DOI:** 10.64898/2025.12.07.25341793

**Authors:** Helen Nguyen, Julie Brown, Lisa Keay

**Affiliations:** Injury Division, The George Institute for Global Health, UNSW Sydney; School of Optometry and Vision Science, UNSW Sydney

**Keywords:** older drivers, licensing, mobility, driving cessation, safety

## Abstract

**Background:** New South Wales (NSW) Australia has long standing age-based licensing, however, little is known about how older drivers and their families and caregivers perceive and navigate this system. This study addresses this gap as part of a comprehensive evaluation examining the impact of the 2008 reforms to aged-based licensing on older drivers, families and caregivers.

**Method:** This is a mixed methods study reporting data from focus group discussions (FGDs) and community-wide quantitative surveys. All FGD transcripts were audio-recorded and transcribed verbatim. Content was inductively coded to create preliminary themes by two independent coders, then deductively coded using the Integrative Public-Policy Acceptance Framework as a guide to create additional themes. The surveys were conducted over the phone with target of 600 older drivers and 600 family members and caregivers in NSW capturing their opinions and experiences with the licensing system.

**Results:** Focus groups were conducted with older drivers (n=34) and families and caregivers (n=15) living in NSW, 608 older drivers and 601 family members completed the phone survey. The older drivers, families and caregivers in both the focus groups and the surveys expressed support for the age-based licensing system and viewed the medical and driving assessments as vital components to complete as they can keep older drivers safe on the roads. However, in the focus groups, the older drivers’ engagement with modified licences was low and families and caregivers wanted more support from general practitioners and the licensing authority. The surveys highlighted the need for better communication between older drivers and the licensing authority. Further, families and caregivers responding to the surveys believed the assessments were more stressful for older drivers than the drivers did themselves.

**Conclusions:** Results highlight the need to improve communication of the licensing regulations to older drivers and families and caregivers to ensure continued support and confidence in navigating the system.

## INTRODUCTION

Australia has an ageing driving population. In New South Wales (NSW), the country’s most populous state, the proportion of drivers aged 60 years and older with a driver’s licence increased from 22% in 2010 to 28% in 2024, with the biggest jump recorded for those aged 80 years and older.(1) With more older drivers on the road, it is vital that there is a licensing system that recognises the changes in both the licence holder profiles and transport needs of older adults. Restrictive and complex age-based licensing policies can negatively impact the mobility and independence of older adults, especially older women who tend to give-up their licences earlier than men.(2) Further, restrictions such as driving at night and on major highways can cause greater disadvantages to older drivers in rural areas compared to those in metropolitan regions.(3) Care must also be taken to ensure that policies in the age-based licensing system do not inadvertently cause more older drivers to give-up driving prematurely which can lead to poorer physical and mental health.(4)

NSW reformed its older drivers licensing system in 2008 to include more flexible options for continued licensing of older drivers and a clearer pathway to driving retirement.

Briefly, the main changes made in 2008 were; (1) lowering the requirement of the annual medical assessment from 80 to 75 years old, (2) changing the frequency of the on-road driving assessments for those aged 85 years and older who want to retain their full, unrestricted licences from annually to biennially, (3) changing the design of the on-road driving assessments from a “full” driving test (i.e. the same one that is completed by those in the graduated learner’s licence system) to a safe driving ability test, (4) allowing older drivers the option of undertaking the on-road driving assessment with an accredited, private, driving instructor instead of the instructors employed by the licensing authority, and (5) introducing a new modified licence option with distance-based restrictions for those aged 85 years and older who do not want to sit the on-road driving assessment.

The NSW age-based licensing system has not been updated since 2008 and is still in effect today. However, little is known about how this system is perceived by the public, particularly older drivers, their families and caregivers, who are the groups most impacted by changes to the system. Looking outside of Australia, a US report examining the older driver licencing procedures in 50 states found most older drivers interviewed were satisfied with the licensing process and had favourable opinions about the medical tests and their frequency.(5) However, support for other assessments during licensure for older drivers was mixed. Whilst the on-road driving assessment was accepted by most older drivers, the written assessment, which is compulsory in several states, was not favoured as it was viewed as not indicative of someone’s safe driving ability and better suited for those who have a history of serious citations and crashes.(5)

Eight years after the licensing reforms in 2008, the NSW government conducted a comprehensive evaluation examining the impacts of these licensing changes on older drivers, families and caregivers and key stakeholders. Focus groups discussions (FGDs), stakeholder interviews, state-wide community panel surveys as well as analyses of state licensing, crash and road injury data from January 2005 to December 2014 were completed. Analyses of licensing and crash rates have been published.(6) Here we report on the perspectives of older drivers, family members and caregivers and how they navigate the NSW older drivers’ licensing system after the 2008 reforms. The focus groups offered in-depth data on the behaviour and attitudes held by the older drivers, families and caregivers. The state-wide telephone surveys were informed by the FGDs and provided a more representative picture of how older drivers, families and caregivers perceive the NSW licensing system.

## METHODS

The qualitative study and the panel telephone surveys were approved by the University of Sydney Human Research Ethics Committee (HREC2016/535).

### Focus Group Discussions

In-person FGDs were conducted with older drivers and family members and caregivers residing in three areas in NSW: Coffs Harbour (19 participants), the Hills District (18 participants), and Canterbury (12 participants). There were 6 focus groups conducted with older drivers and 4 focus groups with family and caregivers. Older drivers could participate if they either held a current NSW driving licence and were still driving or had recently stopped driving at the time of recruitment. All families and caregivers in the focus groups were at least 45 years old and either had an older family member or were caring for an older individual who was a driver or had recently stopped driving.

Recruitment was through presentations to community services, councils and senior groups, and flyers in key locations such as community centres, shopping malls and government services offices. Snowball sampling was used to recruit additional participants.

All focus groups were conducted by two trained research team members who used a discussion guide that asked open-ended questions on the awareness, norms, and impact and attitudes the participants may have on the age-based licensing system and other facilitating factors such as alternative transport, consequences of cognitive decline, and impact of driving on quality of life. All focus groups were audio-recorded and transcribed verbatim. All transcripts were imported into QSR International NVivo V.12 qualitative data analysis software to be coded independently. All transcripts were first coded inductively by two researchers independently to create preliminary themes.

These themes were used to inform the final content of the community telephone surveys. The transcripts were then re-coded deductively by a third researcher using components of the Integrative Public-Policy-Acceptance Framework to identify themes specific to the aim of this manuscript. This framework was chosen as it provides a holistic view of why the public may or may not accept certain policies by integrating key psychological determinants, such as trust and fairness, with desire for government support as the key motivational component.(7) All themes from both the inductive and deductive coding steps were then combined and reviewed by all authors before finalisation. Results from the qualitative study are reported according to the Consolidated criteria for reporting qualitative research guidelines.(8)

### Panel Survey

The statewide community telephone survey of at least 600 older drivers aged 65 years and older was conducted by the Survey Research Centre at Edith Cowan University from October – November 2016. The survey collected basic demographic and licensing status and asked questions on their driving space, opinions and experiences with the NSW older drivers’ licensing system, plans for driving retirement and overall quality of life in terms of mobility and independence. Quotas were set by age group (at least 200 each for 65-74 years, 75-85 years, 85 years and older), gender (equal split), and urbanisation (at least 300 urban, 200 regional, and 100 remote) as classified by the Accessibility/Remoteness Index of Australia was set. Similarly, the state-wide community telephone survey of 600 family members or carers of drivers aged 75 years and older was conducted by the Survey Research Centre at Edith Cowan University from November – December 2016. The survey included questions on demographics, the licensing status of the older adult in their family or under their care, their opinions on the licensing system and how it impacts the older adult, and their role in taking care of the older adult and how it effects their personal lives. Selection of family members and caregivers were based upon the age of the older adult either in their family or under their care and urbanisation which had the same quotas set as the older driver survey respondents.

### Statistical Analysis

Summary tables of participant characteristics by age and residential location were produced. Descriptive statistics summarised the continuous variables while frequency counts and percentages of subjects within each category were used to summarise the categorical data. Comparisons between the groups were conducted using independent T tests for continuous data and tests of proportions for categorical data.

## RESULTS

### Older Drivers

#### Focus Group Discussions

Three major themes emerged from the FGDs with older drivers.

1 Older drivers’ support age-based licensing, but unclear communication can cause confusion. (Integrative Public Policy Acceptance Framework components “problem awareness, “support seeking characteristics”, and “desire for government support”)

Most of the participants supported the medical and driving assessments, and accepted these tests as important components of the licensing system which can help individuals gauge whether they can still safely drive or not. Further, most older drivers in each group were aware of possible changes to their driving skills due to age (Table 1, quotes 1-2).

**Table 1.** Themes identified from the focus groups with older drivers and their corresponding framework components and quotes.

| Theme | Theme Description | Public Policy Acceptance Framework components | Quotes |
| --- | --- | --- | --- |
| Older drivers' support age-based licensing, but unclear communication can cause confusion. | Most older drivers supported the medical and driving assessments and viewed them as beneficial to maintaining safety. However, there is confusion around what the assessments are testing and how modified licences are set. | Problem awareness, support seeking characteristics, policy quality and policy compliance. | 1. "I agree with the assessments because you're endangering other lives as well, sometimes you think, you think you're okay but somebody else needs to confirm that." [Group 4; Participant 3].<br>2. " – but we're not as fit as we used to be, and I don't think it's a bad thing to have people looked at, but its not restricting the licence, it's just having a medical at 75." [Group 5, Participant 4].<br>3. "He got his licences at the end of the war, so he didn't even have to go for a test then. So suddenly being tested when you're 85, it's a bit stressful, and he didn't like the stress." [Group 6, Participant 3].<br>4. "... they can get you into a state where you do stupid things just out of fear for what's wrong, especially when you're getting older." [Group 5; Participant 2] |
|  |  |  | 5. "Can I ask a question? Who determines the distance on that modified licence?" [Group 1, Participant 3]<br>6. "I didn't realise it was negotiable. Because I've got a friend who lives down the South Coast who's 90 and she was a bit worried about what would happen if she went again for her licence. So I must tell her that, because that was very restrictive." [Group 2, Participant 3].<br>7. "I've heard different kilometres mentioned by different people. Is there some sort kilometre distance?" [Group 6; Participant 2].<br>8. "I wasn't quite sure of the ages, but I knew at some stage you had to go and get – you know, see a doctor and whatever, and then you'd have to do a driving test and all that sort of thing. But, yeah, I don't know." [Group 4, Participant 9].<br>9. "It's not until 80 you have to have the [driving] test, so, I think that's reasonable." [Group 5, Participant 4].<br>10. "As I said, I don't understand to what extent the test, what do they measure? I have no idea what they<br>are looking for?" [Group 2, Participant 1] |
| Age-based licensing is imperfect but needed because not all unsafe drivers stop by themselves. | Strict assessments are needed to help regulate unsafe drivers who do not want to stop or are unable to self-assess their skills and know when to retire from driving. However, whether the medical assessment and the health professionals who complete it can identify individuals who may be unsafe on the roads, especially those with cognitive decline, remains uncertain. | Support seeking characteristics, desire for government support, policy quality and policy acceptance. | 11. "There are some changes that happen gradually, they may not be perceptible that you've got to any critical point of incapacity. That may not be apparent to you, until after the accidents." [Group 2, Participant 1].<br>12. "You're never going to stop the occasional driver who shouldn't be driving, getting behind the wheel, so I'm in favour of fairly strict testing, because I think you've got to try and pick up" [Group 3 Participant 3].<br>13. "It's got nothing to do with your driving skills at all, it's just a medical report on your health." [Group 3, Participant 4].<br>14. "... reaction time is what was needed because that's what you use when you're driving and that's what its supposed to be about. Not whether you can count backwards in threes or nines or sevens or whatever." [Group 5, Participant 5].<br>15. "I don't know that it's doing a lot, really, to keep people that perhaps are a bit doubtful off the road." [Group 6, Participant 2] |
|  |  |  | 16. “We’ve all done cognitive tests in different places at different times. But the idea of establishing a threshold with something like that is very difficult. I don’t think science has got it right yet.” [Group 6, Participant 4]<br>17. “But that’s not the part that really is the concern. It’s the driving test, I think, which is the hard thing. And if you’re unwell, chances are you’re not going to pass.” [Group 6, Participant 1].<br>18. “This has been foisted on them as something they have to do. Another form to fill out, okay. Tick this, tick that.” [Group 5, Participant 4].<br>19. “He said “I’m more concerned about me standing, dealing with a wheelchair, than I am concerned with you sitting and driving,” which was interesting .... It’s a bit difficult to say as they don’t take their responsibility as seriously as they might, but I suspect that might be the case.” [Group 3, Participant 4].<br>20. “I know doctors, that I don’t go to, who are friends, that I wouldn’t think would be able to make those assessments. They firstly, some<br>people in particular, they lack the skills to do it. But also, wouldn't want to face up and tell somebody "I'm doing to take your licence off you.""<br>[Group 6, Participant 1]. |
| Age should not be the only criteria as everyone "ages" differently. | Age might be the "easiest" criterion currently available, but it may not be the most appropriate as health differs considerable between individuals. The regulations risk coming off as ageist if age continues to be used to decide when assessments are needed. | Desire for government support, policy quality and policy acceptance. | 21. *"They might all be the same age but some people are really on the ball where others mightn't be."* [Group 2, Participant 1].<br>22. *"It shouldn't be any particular age because everybody isn't the same at that age are they. Some people are more slower and some not."* [Group 1, Participant 3].<br>23. *"There's such a variation in people, I mean, I suppose you've just got to find a good average age for it. There's obviously maybe somebody at 70 who needs a medical and somebody at 85 who may not need a medical so I suppose you've got to pick a happy medium."* [Group 4, Participant 5]<br>24. *"There's a lot younger that aren't really on the ball that much. I think there should be more tests for the younger ones periodically."* [Group 1, Participant 2].<br>25. *"It would be some question whether 75 is not even too late to start medical assessments because, you*<br>*know, people with heart conditions and so on, that can come into play long before they're 75."* [Group 5, Participant 1].<br>26. *"Well I don't know whether you can make other laws for older drivers. Laws apply to everybody. Irrespective of age. From a P-plater to an old bastard, 85. Nowadays, there are still people driving out there who are in their 90s. So there's a law for everybody."* [Group 4, Participant 4]<br>27. *"I think there should be a uniform law right across Australia so everybody – we know that, you know, there are safeguards."* [Group 5, Participant 5] |

However, even though some participants felt that the changes to the on-road driving test made it more *“user friendly” [Group 2, Participant 1]* and *“good…Very intensive” [Group 5, Participant 1]*, others found it to be stressful. This anxiety may be because many had not taken driving tests to receive or retain their licence so being asked to sit one after driving for most of their lives is a foreign concept. The added pressure of potentially failing the test and not being able to renew their licence can also induce stress, especially amongst those who are not confident drivers to begin with (Table 1, quotes 3-4).

When prompted by the facilitator that modified licences were an option for those uncomfortable with taking the driving assessment, confusion on how to obtain a modified licence and how its distance-based restrictions were set, was raised in all focus groups, thereby showing that engagement with rules on modified licences is low (Table 1, quotes 5-7).

There was also notable confusion on when older drivers are expected to complete their medical and on-road driving assessments. Some participants either did not know or incorrectly quoted the age they thought the tests would be required. Others also expressed confusion around what these tests would involve. These misunderstandings, while easily addressed, reveal how communication on all components of the licensing system licence changes may not be effectively reaching older drivers (Table 1, quotes 8-10).

2 Age-based licensing is imperfect but needed because not all unsafe drivers stop by themselves. (Integrative Public-Policy Acceptance Framework components “problem awareness”, “support-seeking characteristics”, “desire for government support” and “policy qualities”)

Participants felt, that for most of the time, rigid medical and driving assessments are needed because many people are incapable of gauging declines in their own driving skills. While the first theme showed that participants were confident in recognising changes in their own driving abilities and using the assessment to validate these perceptions, the second theme reveals that their support for the assessment is also based on the belief that there are older drivers in the community who will not self-regulate unless they are required to either by law following a crash(Table 1, quotes 11-12).

There were mixed views on how well they thought the medical tests appropriately identifies and assesses those with cognitive decline. Some felt that it is neither scientifically robust enough nor applicable as it does not test skills that relate to safe driving (Table 1, quotes 13-16). However, few participants did voice that if the medical assessment was unable to identify someone with cognitive decline, then the driving test is stringent enough to fill in the gap (Table 1, quote 17).

Participants also held reservations about a general practitioner’s ability to appropriately test and link cognitive decline to safe driving ability. Even though some participants did describe their doctors completing the medical assessment thoroughly and conscientiously, the majority believed general practitioners to place little faith in these assessments. This is because they felt general practitioners were already responsible for many different things which therefore meant they tended to place more concern on other health conditions. Older drivers also pointed out that some general practitioners may feel pressured to be less than truthful when completing the medical assessment due to concerns that their patient-doctor relationship may be negatively impacted, especially if the older adult has been their patient for several years (Table 1, quotes 18-20).

3 Age should not be the only criteria as everyone “ages” differently. (Integrative Public-Policy Acceptance Framework components “policy qualities” and “policy acceptance”)

The belief that age is not a good indicator of an individual’s ability to safely drive was brought up in all focus groups. Health varies between individuals regardless of their age. This is why participants voiced concerns on medical testing beginning at 75 years, as some in the community may need to complete it earlier and others not until much later. They also pointed out the reliance on age as a criterion may also be why picking up cognitive decline is so difficult and why some individuals are able to retain their licence even though their health may negatively impact their driving skills (Table 1, quotes 21-23).

This is why a concept emerged that the rules discriminated against older drivers. They expressed that the system should be made fairer by either lowering the age of testing, especially the medical assessment, or introducing similar licensing laws to drivers of all age groups. Some participants also felt that the system could be safer if the driving assessment was completed annually as *“a lot can happen in two years” [Group 4, Participant 2]* because *“after 85 years, your reaction time is sort of diminishing at a very sharp angle” [Group 5, Participant 3].* While the older drivers recognised that these changes would require select individuals in the community to complete assessments earlier and more regularly, they accepted this as they viewed the measures as a potential way to create a safer and fairer road system for all (Table 1, quotes 24-27).

### Community Surveys

A total of 608 older drivers (52% females) completed the telephone survey. The majority held a full licence (88.3%), only drove a car (89.4%) and never used alternative transport (42.9%). Supplementary Table 1 details the characteristics of the older drivers surveyed.

The beliefs described in the themes from the qualitative study were supported by the majority (Table 3). Most respondents agreed that the medical assessment was a fair way of determining licensing (81%) and that it would help improve safety for all on the roads (75%). However, when split by age group, there were more disagreement on both fairness (p=0.0164) and safety benefit (p<0.0001) among the younger age groups (65-84 years) compared to the oldest age group (85 years and older). More drivers living in remote areas held “neutral” opinions on the safety benefit of the medical assessment, but the overall majority (68%) still agreed that the assessment would likely help improve road safety for all. Support for the on-road practical assessment was even higher with 80% and 90% agreeing that the assessment would improve safety for everyone on the road and was a fair way to determine licensing for people aged 85 years and over, respectively. There were no significant differences in the opinions held on the on-road assessment by age groups or location of residence.

The survey respondents, regardless of their age and area of residence, believed age and not driving skills, should determine licensing with 89% disagreeing with using skill over age. This may be why 69% agreed that starting the medical assessment at 75 years was appropriate and that 70% felt that completing the driving test every two years was “about right”. Further, even though 66% of those who had completed the driving test did not feel anxious about it at all, 33% indicated some level of anxiousness.

When looking at those aged 74 years and older who had received a letter from the licencing authority about the medical test, 70% (213/304) agreed that the letter provided helpful and relevant information about their upcoming medical assessment (Figure 1). Similarly, 72% (95/132) of those aged 84 years and older found the letter about their upcoming driving test to be helpful (Figure 1). However, these letters were not being received by all older drivers. Only 72% (304/425) reported ever receiving a letter about the medical assessment and even fewer (63% (132/209)) had received any communication about the driving test. Older drivers living in urban, rural, or remote areas were equally likely to report not receiving either letter.

**Figure 1.**
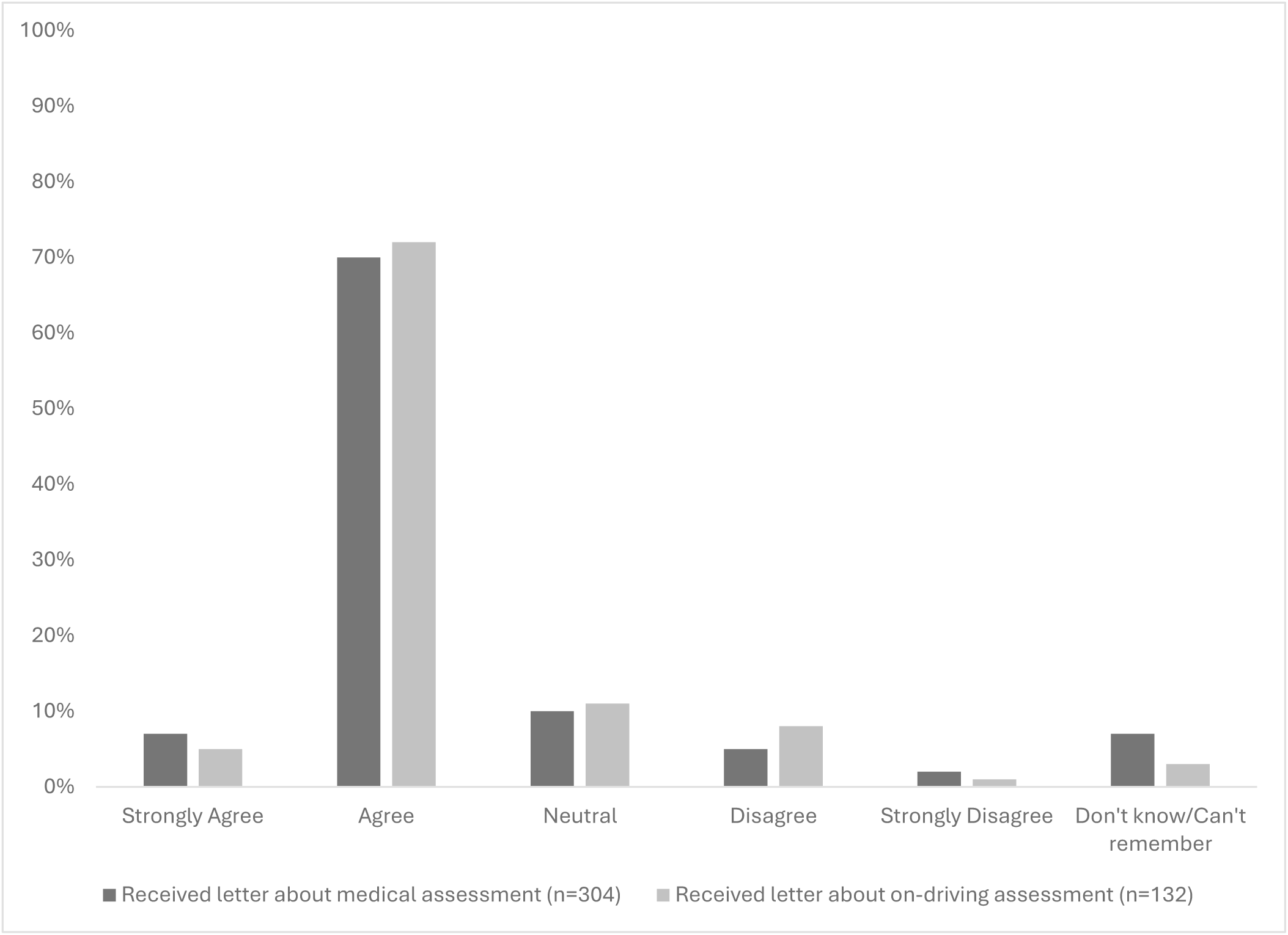
The extent to which older drivers agree that the letters they receive from the government about their annual medical assessment and on-road driving assessment are helpful

### Family members and caregivers

#### Focus Group Discussions

Two major themes emerged from the focus groups with family members and caregivers.

1 Recognition that family and caregivers contribute to implementation of the age-based licensing system. (Integrative Public-Policy Acceptance Framework components “problem awareness”, “support-seeking characteristics” and “desire for government support”)

All family members and caregivers understood that the regulations of the age-based licensing system were created to keep older adults safe on the roads. They therefore felt that they play a major role in creating a safer road environment by helping older drivers navigate the licensing process and addressing their mobility needs after driving retirement. Family members and caregivers intervened to make sure that the older driver stopped driving if they had concerns for their safety and provided tips on how to pass the on-road driving assessment if the older adult would like to keep their unrestricted licence (Table 2, quote 1-2). However, some did find it difficult to intervene as approaching the topic of licensing incorrectly can lead to relationship breakdown (Table 2, quote 3).

**Table 2.** Themes identified from the focus groups with family members and caregivers and their corresponding framework components and quotes.

| Theme | Theme Description | Public Policy Acceptance Framework components | Quotes |
| --- | --- | --- | --- |
| Recognition that family and caregivers contribute to implementation of the age-based licensing system. | Families and caregivers help older drivers navigate the licensing system and their post-driving mobility needs. It is therefore crucial that they are well-supported to minimise any undue burden their responsibility might entail. | Problem awareness, desire for government support and policy compliance. | 1. “They have been to the doctor’s as in they my sister and her husband and the family and everyone, begging them to take his licence off. They’ve been to the licensing people saying please take his licence off him.” [Group 3, Participant 4]<br>2. “I’ll try and talk him around and try and explain to him the rules of roundabouts because that was my biggest concern with him...” [Group 2, Participant 2].<br>3. “... like in your case, you could have had a mother that disowned you at 90 something all because she lost her licence.” [Group 3, Participant 1].<br>4. “I don’t think we have any choice. I think we just take this as being something like sunshine and sundown and we have to live with it.” [Group 1, Participant 2].<br>5. “... someone has to step in, and that’s usually family, but that creates another whole lot of issues.” [Group 3, Participant 3] |
|  |  |  | 6. “Yeah I think you would obviously put yourself at their beck and – well, no, I won’t say at their beck and call. But if there was something – I mean i’m thinking of two people that are at opposite ends of the state here but if they were in Sydney, you know, I would feel like I had to go and get my aunty and take her to her appointment...” [Group 4, Participant 3]<br>7. “I think, though, people should work together to do it. I just really don’t think – I wouldn’t like to let my daughter loose on my quite honestly. I’d be off the road now but, yeah, family working in conjunction with – any medical.” [Group 4, Participant 1].<br>8. “Maybe the RMS becoming a bit of a referral base as well. So not just going, “You can’t drive anymore or you can only drive in these areas” they could say, “Look, these are the areas you. If you need some support, here, I’ll refer you on to dot dot dot.” [Group 3, Participant 2]. |
| Desire for evidence and a clear rationale for age-based licensing. | Increasing evidence on how the age-based licensing system impacts safety for older drivers may clear up misunderstandings and lead to | Policy quality, policy acceptance and policy compliance. | 9. “I think statistics need to have shown that its better, worse or indifferent” [Group 1, Participant 2]<br>10. “It would be interesting to see the evidence because for all the reasons we |
|  | continued acceptance in the community. |  | talked about before when older drivers don't tend to driver as far and tend to be a bit more cautious. I don't know if they really do represent the risk we perceive to be the risk." [Group 1, Participant 3].<br>11. "You see one person, one older person has an accident and then all hell breaks loose, but every night you listen to the news, there's two or three dill brained young people under 30." [Group 1, Participant 1]<br>12. "And I think that was a bit of a knee jerk reaction and sure there's older drivers that do do some silly things like putting your foot on the accelerator instead of the brake but, God, young people do it too." [Group 4, Participant 2]. |

**Table 3.** Views and opinions on licencing system from older drivers stratified by location and age group (n=608)

| Question | Option | Location |  |  |  | P-value* | Age group, years |  |  |  |  |
| --- | --- | --- | --- | --- | --- | --- | --- | --- | --- | --- | --- |
|  |  | Urban<br>(n=303,<br>%) | Rural<br>(n=203,<br>%) | Remote<br>(n=102,<br>%) | Total (n=608,<br>%) |  | 65-74<br>(n=209,<br>%) | 75-84<br>(n=199,<br>%) | 85+<br>(n=200,<br>%) | Total<br>(n=608, %) | P-value* |
| A medical examination every year is a fair way to determine licensing for older drivers | Strongly Disagree | 4 (1) | 7 (3) | 4 (4) | 15 (2) | 0.05 | 9 (4) | 4 (2) | 2 (1) | 15 (2) | 0.0164 |
|  | Disagree | 31 (10) | 20 (10) | 6 (6) | 57 (9) |  | 30 (14) | 16 (8) | 11 (6) | 57 (9) |  |
|  | Neutral | 17 (6) | 12 (6) | 13 (13) | 42 (7) |  | 15 (7) | 17 (9) | 10 (5) | 42 (7) |  |
|  | Agree | 176 (58) | 124 (61) | 64 (63) | 364 (60) |  | 114 (55) | 122 (61) | 128 (64) | 364 (60) |  |
|  | Strongly Agree | 75 (25) | 40 (20) | 15 (15) | 130 (21) |  | 41 (20) | 40 (20) | 49 (25) | 130 (21) |  |
| A medical examination every year is likely to improve safety for everyone on the road | Strongly Disagree | 11 (4) | 6 (3) | 2 (2) | 19 (3) | 0.0226 | 13 (6) | 5 (3) | 1 (1) | 19 (3) | <0.0001 |
|  | Disagree | 51 (17) | 23 (11) | 14 (14) | 88 (14) |  | 42 (20) | 32 (16) | 14 (7) | 88 (14) |  |
|  | Neutral | 17 (6) | 14 (7) | 17 (17) | 48 (8) |  | 16 (8) | 18 (9) | 14 (7) | 48 (8) |  |
|  | Agree | 163 (54) | 124 (61) | 51 (50) | 338 (56) |  | 92 (44) | 110 (55) | 136 (68) | 338 (56) |  |
|  | Strongly Agree | 61 (20) | 36 (18) | 18 (18) | 115 (19) |  | 46 (22) | 34 (17) | 35 (18) | 115 (19) |  |
| 75 is the right age to begin having a medical examination every year to keep your driver's licence | Strongly Disagree | 14 (5) | 4 (2) | 1 (1) | 19 (3) | 0.70 | 9 (4) | 7 (4) | 3 (2) | 19 (3) | 0.40 |
|  | Disagree | 45 (15) | 32 (16) | 17 (17) | 94 (15) |  | 34 (16) | 35 (18) | 25 (13) | 94 (15) |  |
|  | Neutral | 37 (12) | 28 (14) | 14 (14) | 79 (13) |  | 25 (12) | 21 (11) | 33 (17) | 79 (13) |  |
|  | Agree | 173 (57) | 120 (59) | 59 (58) | 352 (58) |  | 121 (58) | 112 (56) | 119 (60) | 352 (58) |  |
|  | Strongly Agree | 34 (11) | 19 (9) | 11 (11) | 64 (11) |  | 20 (10) | 24 (12) | 20 (10) | 64 (11) |  |
| A modified licence is a good in-between step from a full licence to retiring from driving | Strongly disagree | 10 (3) | 11 (5) | 2 (2) | 23 (4) | 0.64 | 8 (4) | 6 (3) | 9 (5) | 23 (4) | 0.16 |
|  | Disagree | 45 (15) | 26 (13) | 10 (10) | 81 (13) |  | 23 (11) | 25 (13) | 33 (17) | 81 (13) |  |
|  | Neutral | 27 (9) | 21 (10) | 14 (14) | 62 (10) |  | 24 (11) | 14 (7) | 24 (12) | 62 (10) |  |
|  | Agree | 175 (58) | 113 (56) | 61 (60) | 349 (57) |  | 118 (56) | 130 (65) | 101 (51) | 349 (57) |  |
|  | Strongly agree | 46 (15) | 32 (16) | 15 (15) | 93 (15) |  | 36 (17) | 24 (12) | 33 (17) | 93 (15) |  |
| A Practical Driving Test for people 85 and over to keep a full licence is likely to improve safety for everyone on the road | Strongly disagree | 4 (1) | 6 (3) | 1 (1) | 11 (2) | 0.22 | 4 (2) | 3 (2) | 4 (2) | 11 (2) | 0.80 |
|  | Disagree | 36 (12) | 20 (10) | 9 (9) | 65 (11) |  | 22 (11) | 23 (12) | 20 (10) | 65 (11) |  |
|  | Neutral | 16 (5) | 12 (6) | 13 (13) | 41 (7) |  | 15 (7) | 13 (7) | 13 (7) | 41 (7) |  |
|  | Agree | 191 (63) | 130 (64) | 65 (64) | 386 (63) |  | 124 (59) | 127 (64) | 135 (68) | 386 (63) |  |
|  | Strongly agree | 56 (18) | 35 (17) | 14 (14) | 105 (17) |  | 44 (21) | 33 (17) | 28 (14) | 105 (17) |  |
| A practical driving test is a fair way to determine licensing among people 85 and over | Strongly disagree | 3 (1) | 3 (1) | 0 (0) | 6 (1) | 0.61 | 0 (0) | 2 (1) | 4 (2) | 6 (1) | 0.37 |
|  | Disagree | 18 (6) | 9 (4) | 8 (8) | 35 (6) |  | 10 (5) | 11 (6) | 14 (7) | 35 (6) |  |
|  | Neutral | 10 (3) | 11 (5) | 4 (4) | 25 (4) |  | 6 (3) | 11 (6) | 8 (4) | 25 (4) |  |
|  | Agree | 222 (73) | 146 (72) | 79 (77) | 447 (74) |  | 154 (74) | 147 (74) | 146 (73) | 447 (74) |  |
|  | Strongly agree | 50 (17) | 34 (17) | 11 (11) | 95 (16) |  | 39 (19) | 28 (14) | 28 (14) | 95 (16) |  |
| Licensing should be determined by skill not age | Strongly disagree | 138 (46) | 82 (40) | 45 (44) | 265 (44) | 0.60 | 89 (43) | 95 (48) | 81 (41) | 265 (44) | 0.38 |
|  | Disagree | 137 (45) | 97 (48) | 42 (41) | 276 (45) |  | 99 (47) | 82 (41) | 95 (48) | 276 (45) |  |
|  | Neutral | 11 (4) | 11 (5) | 6 (6) | 28 (5) |  | 10 (5) | 7 (4) | 11 (6) | 28 (5) |  |
|  | Agree | 13 (4) | 8 (4) | 8 (8) | 29 (5) |  | 10 (5) | 12 (6) | 7 (4) | 29 (5) |  |
|  | Strongly agree | 4 (1) | 5 (2) | 1 (1) | 10 (2) |  | 1 (0) | 3 (2) | 6 (3) | 10 (2) |  |
| For Drivers 85 years and over, do you think that a driving test every two years is: | Too Often | 18 (6) | 14 (7) | 5 (5) | 37 (6) | 0.60 | 4 (2) | 18 (9) | 15 (8) | 37 (6) | 0.0011 |
|  | About Right | 212 (70) | 146 (72) | 70 (69) | 428 (70) |  | 143 (68) | 137 (69) | 148 (74) | 428 (70) |  |
|  | Not often enough | 59 (19) | 33 (16) | 25 (25) | 117 (19) |  | 55 (26) | 37 (19) | 25 (13) | 117 (19) |  |
|  | Don't know/Not sure | 14 (5) | 10 (5) | 2 (2) | 26 (4) |  | 7 (3) | 7 (4) | 12 (6) | 26 (4) |  |

\* Presented as 2 decimal places unless significant (<0.05).

Some however, felt the responsibility of keeping older adults, who are either transitioning to driving retirement or no longer drive, active and mobile was too reliant on the contributions from family and caregivers alone. The little support from other sectors, such as licensing authorities and health, can cause a strain on family relationships. The major cause of this resentment is because family members and caregivers felt like they had little to no choice but to adjust their lifestyles to now accommodate for the older adult. In areas with poor public transport and alternative transport options, supporting their older adult could *“curtail some of my social activities” [Group 4, Participant 2]* and *“cause major family upheavals” [Group 3, Participant 1].* The extent to which older adults rely on family and caregivers also depended on their relationship and not all individuals are comfortable with or able to stay in contact (Table 2, quotes 4-6).

There was therefore a desire to have more engagement from general practitioners and the licensing authorities to create a more holistic licensing system where the onus of keeping older adults safe and mobile is more evenly shared. (Table 2, quotes 7-8)

2 Desire for evidence and a clear rational for age-based licensing (Integrative Public-Policy Acceptance Framework components “policy qualities” and “policy acceptance”)

All participants mentioned that while they supported the age-based licensing system, they would like more transparency on evidence showing how these laws have impacted safety for older drivers. Families and caregivers who were close to undergoing the assessments themselves were especially vocal about this matter. Participants believed evidence would help determine whether acceptance and support of the licensing system would continue or not (Table 2, quotes 9-10).

Because there is also a lack of information on how the age-based licensing laws were created, some participants felt that the rules may be prejudiced against older drivers. Public condemnation may have fuelled the creation of the system therefore making older drivers more targeted than younger-aged drivers who commit the same errors (Table 2, quotes 11-12).

#### Community Surveys

The telephone surveys were completed by 601 family members and caregivers (62.8% females) either related to or looking after 602 older adults (Supplementary Table 2).

Most of the older adults were male (55%) and had a car licence (94.4%).

Like the surveyed older drivers, support for the medical and on-road driving assessments was high among surveyed family and carers (Table 4): 73% believed the medical assessment was a fair way to determine licensing while 67% thought it would lead to improvements in safety for all on the road - a notably smaller proportion compared to the surveyed older drivers. There were no differences in agreement when responses were split by their area of residence or age group of the older adult they were related to or caring for. Support for the driving test was higher with 88% agreeing that the assessment was a fair way of determining licensing and 83% agreed that the driving test would lead to improvement in safety for all. Having the driving test every two years was also considered “about right” by 62% with the highest agreement coming from those who live in remote areas. Further, families and caregivers also disagreed with skill being used to determine licensing (82%).

**Table 4.** Views and opinions of licencing from family and caregivers stratified by location and the age group of the older driver in their family or that they care for (n=602)

| Question | Option | Location |  |  |  | P-value* | Age group, years |  |  |  | P-value* |
| --- | --- | --- | --- | --- | --- | --- | --- | --- | --- | --- | --- |
|  |  | Urban (n=298, %) | Rural (n=240, %) | Remote (n=64, %) | Total (n=602, %) |  | 65-74 (n=240, %) | 75-84 (n=201, %) | 85+ (n=161, %) | Total (n=602, %) |  |
| A medical examination every year is a fair way to determine licensing for older drivers | Strongly Disagree | 8 (3) | 13 (5) | 2 (3) | 23 (4) | 0.15 | 7 (3) | 7 (3) | 9 (6) | 23 (4) | 0.11 |
|  | Disagree | 47 (16) | 20 (8) | 11 (17) | 78 (13) |  | 43 (18) | 21 (10) | 14 (9) | 78 (13) |  |
|  | Neutral | 25 (8) | 28 (12) | 5 (8) | 58 (10) |  | 20 (8) | 18 (9) | 20 (12) | 58 (10) |  |
|  | Agree | 140 (47) | 122 (51) | 29 (45) | 291 (48) |  | 110 (46) | 106 (53) | 75 (47) | 291 (48) |  |
|  | Strongly Agree | 78 (26) | 57 (24) | 17 (27) | 152 (25) |  | 60 (25) | 49 (24) | 43 (27) | 152 (25) |  |
| A medical examination every year is likely to improve safety for everyone on the road | Strongly Disagree | 13 (4) | 10 (4) | 3 (5) | 26 (4) | 0.71 | 12 (5) | 9 (4) | 5 (3) | 26 (4) | 0.94 |
|  | Disagree | 51 (17) | 42 (18) | 12 (19) | 105 (17) |  | 41 (17) | 32 (16) | 32 (20) | 105 (17) |  |
|  | Neutral | 27 (9) | 34 (14) | 4 (6) | 65 (11) |  | 25 (10) | 25 (12) | 15 (9) | 65 (11) |  |
|  | Agree | 144 (48) | 109 (45) | 32 (50) | 285 (47) |  | 113 (47) | 93 (46) | 79 (49) | 285 (47) |  |
|  | Strongly Agree | 63 (21) | 45 (19) | 13 (20) | 121 (20) |  | 49 (20) | 42 (21) | 30 (19) | 121 (20) |  |
| 75 is the right age to begin having a medical examination every year to keep a driver's licence | Strongly Disagree | 16 (5) | 13 (5) | 3 (5) | 32 (5) | 0.89 | 15 (6) | 7 (3) | 10 (6) | 32 (5) | 0.71 |
|  | Disagree | 58 (19) | 40 (17) | 13 (20) | 111 (18) |  | 44 (18) | 34 (17) | 33 (21) | 111 (18) |  |
|  | Neutral | 39 (13) | 34 (14) | 11 (17) | 84 (14) |  | 31 (13) | 34 (17) | 19 (12) | 84 (14) |  |
|  | Agree | 140 (47) | 124 (52) | 27 (42) | 291 (48) |  | 116 (48) | 101 (50) | 74 (46) | 291 (48) |  |
|  | Strongly Agree | 45 (15) | 29 (12) | 10 (16) | 84 (14) |  | 34 (14) | 25 (12) | 25 (16) | 84 (14) |  |
| A modified licence is a good in-between step from a | Strongly Disagree | 11 (4) | 6 (3) | 2 (3) | 19 (3) | 0.44 | 8 (3) | 5 (2) | 6 (4) | 19 (3) | 0.82 |
|  | Disagree | 23 (8) | 8 (3) | 3 (5) | 34 (6) |  | 14 (6) | 13 (6) | 7 (4) | 34 (6) |  |
|  | Neutral | 18 (6) | 17 (7) | 5 (8) | 40 (7) |  | 19 (8) | 12 (6) | 9 (6) | 40 (7) |  |
|  | Agree | 148 (50) | 139 (58) | 33 (52) | 320 (53) |  | 131 (55) | 108 (54) | 81 (50) | 320 (53) |  |
| full licence to retiring from driving | Strongly Agree | 98 (33) | 70 (29) | 21 (33) | 189 (31) |  | 68 (28) | 63 (31) | 58 (36) | 189 (31) |  |
| A practical driving test for people 85 years and over to keep a full licence is likely to improve safety for everyone on the road | Strongly Disagree | 7 (2) | 8 (3) | 0 (0) | 15 (2) | 0.34 | 8 (3) | 4 (2) | 3 (2) | 15 (2) | 0.61 |
|  | Disagree | 27 (9) | 16 (7) | 4 (6) | 47 (8) |  | 16 (7) | 17 (8) | 14 (9) | 47 (8) |  |
|  | Neutral | 24 (8) | 14 (6) | 2 (3) | 40 (7) |  | 21 (9) | 12 (6) | 7 (4) | 40 (7) |  |
|  | Agree | 146 (49) | 137 (57) | 39 (61) | 322 (53) |  | 122 (51) | 114 (57) | 86 (53) | 322 (53) |  |
|  | Strongly Agree | 94 (32) | 65 (27) | 19 (30) | 178 (30) |  | 73 (30) | 54 (27) | 51 (32) | 178 (30) |  |
| A practical driving test is a fair way to determine licensing among people 85 and over | Strongly Disagree | 8 (3) | 1 (0) | 1 (2) | 10 (2) | 0.44 | 7 (3) | 2 (1) | 1 (1) | 10 (2) | 0.17 |
|  | Disagree | 19 (6) | 9 (4) | 3 (5) | 31 (5) |  | 10 (4) | 15 (7) | 6 (4) | 31 (5) |  |
|  | Neutral | 13 (4) | 15 (6) | 4 (6) | 32 (5) |  | 13 (5) | 9 (4) | 10 (6) | 32 (5) |  |
|  | Agree | 183 (61) | 153 (64) | 43 (67) | 379 (63) |  | 146 (61) | 135 (67) | 98 (61) | 379 (63) |  |
|  | Strongly Agree | 75 (25) | 62 (26) | 13 (20) | 150 (25) |  | 64 (27) | 40 (20) | 46 (29) | 150 (25) |  |
| Licensing should be determined by skill not age | Strongly Disagree | 120 (40) | 113 (47) | 34 (53) | 267 (44) | 0.30 | 97 (40) | 89 (44) | 81 (50) | 267 (44) | 0.60 |
|  | Disagree | 117 (39) | 94 (39) | 19 (30) | 230 (38) |  | 93 (39) | 79 (39) | 58 (36) | 230 (38) |  |
|  | Neutral | 13 (4) | 9 (4) | 2 (3) | 24 (4) |  | 12 (5) | 7 (3) | 5 (3) | 24 (4) |  |
|  | Agree | 23 (8) | 10 (4) | 6 (9) | 39 (6) |  | 17 (7) | 12 (6) | 10 (6) | 39 (6) |  |
|  | Strongly Agree | 25 (8) | 14 (6) | 3 (5) | 42 (7) |  | 21 (9) | 14 (7) | 7 (4) | 42 (7) |  |
| For drivers 85 years and over, do you think that a driving test every two years is: | Too Often | 15 (5) | 5 (2) | 3 (5) | 23 (4) | 0.26 | 11 (5) | 10 (5) | 2 (1) | 23 (4) | 0.48 |
|  | About Right | 175 (59) | 154 (64) | 43 (67) | 372 (62) |  | 144 (60) | 124 (62) | 104 (65) | 372 (62) |  |
|  | Not often enough | 97 (33) | 68 (28) | 14 (22) | 179 (30) |  | 71 (30) | 60 (30) | 48 (30) | 179 (30) |  |
|  | Don't know/Not Sure | 11 (4) | 13 (5) | 4 (6) | 28 (5) |  | 14 (6) | 7 (3) | 7 (4) | 28 (5) |  |

\* Presented as 2 decimal places unless significant (<0.05).

Unlike the surveyed older adults, family members and caregivers tended to think older drivers were more nervous about the medical and driving tests than the older drivers themselves (Figure 2: 88% (530/604) of older drivers felt “not at all” nervous about completing the annual medical assessment to keep their licence while only 23% (138/602) of family and caregivers felt the older adult was not nervous. Similarly, only 5% (14/311) of older drivers reported feeling nervous about taking the practical driving test compared to 26% (30/116) of the family and caregivers who felt that the older driver was nervous before the test.

**Figure 2.**
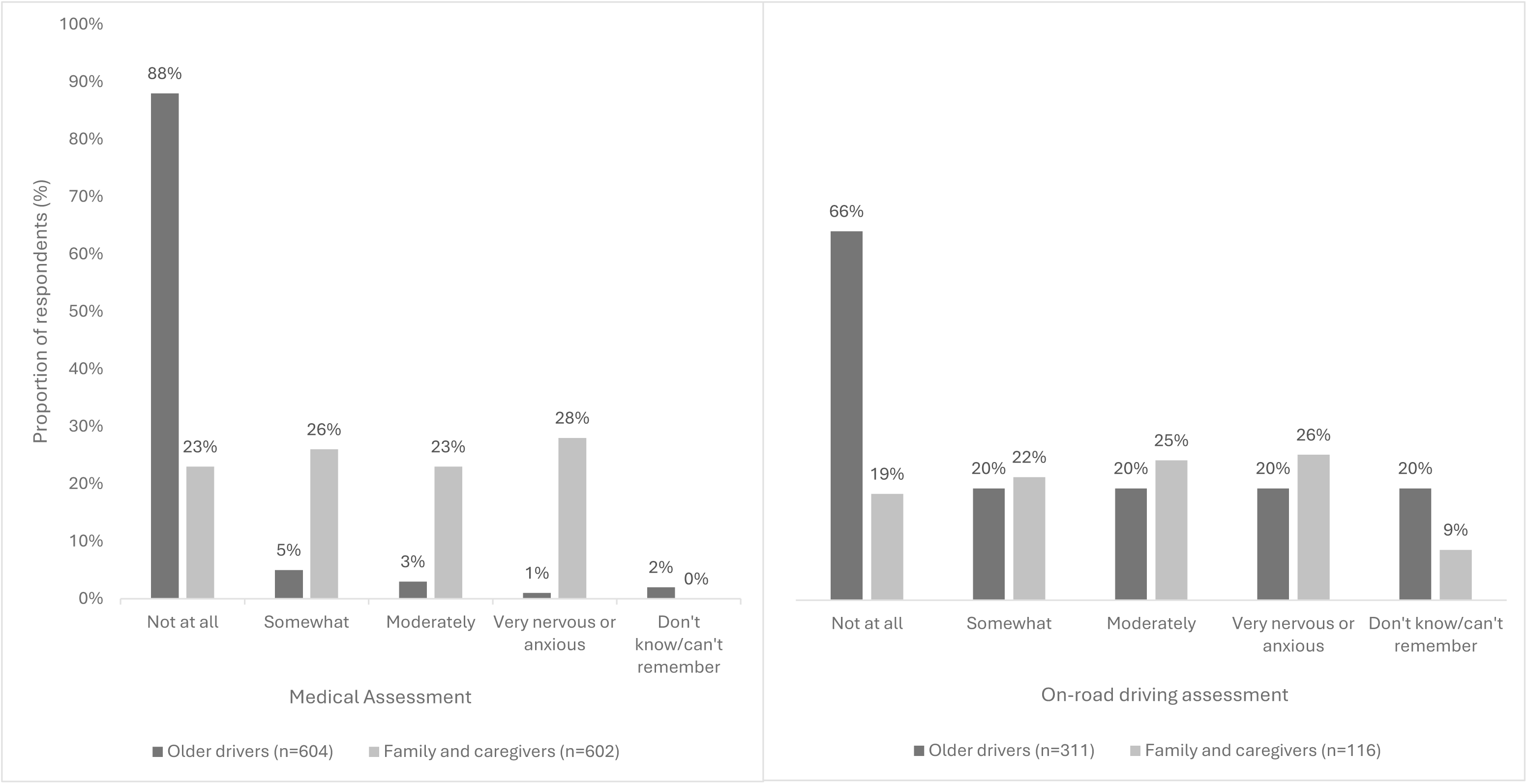
Level of anxiety older drivers, family members and caregivers have on completing the annual medical assessment (left) and on-road driving assessment (right)

The surveyed family members and caregivers were less varied on their opinions regarding how burdensome they felt it was to provide transport for the older person (Table 5). The majority found it rewarding to provide transport for the older driver (64%) and did not find providing transport to conflict with their work schedule (77%). However, there was more variation in responses on work conflict from those living in remote areas than urban and rural (p=0.0042). Providing transport for the older driver was viewed as a rewarding experience (p=0.0304) that did not disrupt social or family time (p=0.0038).

**Table 5.** Views and opinions from family and caregivers stratified about providing transport to older adults in the last 6 months (n=377)

| Question | Options | Location |  |  |  |  | Age groups, years |  |  |  |  |
| --- | --- | --- | --- | --- | --- | --- | --- | --- | --- | --- | --- |
|  |  | Urban<br>(n=298,<br>%) | Rural<br>(n=240,<br>%) | Remote<br>(n=64, %) | Total (n=602,<br>%) | P-value* | 65-74<br>(n=240,<br>%) | 75-84<br>(n=201,<br>%) | 85+<br>(n=161,<br>%) | Total<br>(n=602,<br>%) | P-value* |
| Providing transport for the older person is stressful for me | Strongly Disagree | 48 (27) | 40 (30) | 5 (18) | 93 (28) | 0.09 | 52 (31) | 18 (19) | 23 (30) | 93 (28) | 0.30 |
|  | Disagree | 95 (54) | 56 (42) | 14 (50) | 165 (49) |  | 81 (48) | 48 (52) | 36 (47) | 165 (49) |  |
|  | Neutral | 12 (7) | 16 (12) | 3 (11) | 31 (9) |  | 13 (8) | 8 (9) | 10 (13) | 31 (9) |  |
|  | Agree | 19 (11) | 16 (12) | 3 (11) | 38 (11) |  | 18 (11) | 15 (16) | 5 (7) | 38 (11) |  |
|  | Strongly Agree | 2 (1) | 5 (4) | 3 (11) | 10 (3) |  | 4 (2) | 4 (4) | 2 (3) | 10 (3) |  |
| Providing transport is time consuming for me | Strongly Disagree | 40 (23) | 33 (25) | 4 (14) | 77 (23) | 0.09 | 44 (26) | 16 (17) | 17 (22) | 77 (23) | 0.18 |
|  | Disagree | 86 (49) | 51 (38) | 11 (39) | 148 (44) |  | 78 (46) | 44 (47) | 26 (34) | 148 (44) |  |
|  | Neutral | 11 (6) | 21 (16) | 3 (11) | 35 (10) |  | 17 (10) | 10 (11) | 8 (11) | 35 (10) |  |
|  | Agree | 36 (20) | 24 (18) | 8 (29) | 68 (20) |  | 27 (16) | 19 (20) | 22 (29) | 68 (20) |  |
|  | Strongly Agree | 3 (2) | 4 (3) | 2 (7) | 9 (3) |  | 2 (1) | 4 (4) | 3 (4) | 9 (3) |  |
| Providing transport often conflicts with my work schedule | Strongly Disagree | 55 (31) | 47 (35) | 3 (11) | 105 (31) | 0.0042 | 66 (39) | 20 (22) | 19 (25) | 105 (31) | 0.0084 |
|  | Disagree | 84 (48) | 60 (45) | 10 (36) | 154 (46) |  | 74 (44) | 48 (52) | 32 (42) | 154 (46) |  |
|  | Neutral | 13 (7) | 14 (11) | 5 (18) | 32 (10) |  | 13 (8) | 10 (11) | 9 (12) | 32 (10) |  |
|  | Agree | 18 (10) | 10 (8) | 9 (32) | 37 (11) |  | 12 (7) | 10 (11) | 15 (20) | 37 (11) |  |
|  | Strongly Agree | 6 (3) | 2 (2) | 1 (4) | 9 (3) |  | 3 (2) | 5 (5) | 1 (1) | 9 (3) |  |
| Providing transport is rewarding for me | Strongly Disagree | 5 (3) | 10 (8) | 0 (0) | 15 (4) | 0.89 | 8 (5) | 3 (3) | 4 (5) | 15 (4) | 0.0304 |
|  | Disagree | 25 (14) | 9 (7) | 2 (7) | 36 (11) |  | 27 (16) | 6 (6) | 3 (4) | 36 (11) |  |
|  | Neutral | 30 (17) | 36 (27) | 8 (29) | 74 (22) |  | 42 (25) | 19 (20) | 13 (17) | 74 (22) |  |
|  | Agree | 96 (55) | 64 (48) | 15 (54) | 175 (52) |  | 75 (45) | 56 (60) | 44 (58) | 175 (52) |  |
|  | Strongly Agree | 20 (11) | 14 (11) | 3 (11) | 37 (11) |  | 16 (10) | 9 (10) | 12 (16) | 37 (11) |  |
| Providing transport takes time away from my social or family life | Strongly Disagree | 49 (28) | 39 (29) | 2 (7) | 90 (27) | 0.0038 | 55 (33) | 17 (18) | 18 (24) | 90 (27) | 0.10 |
|  | Disagree | 93 (53) | 64 (48) | 14 (50) | 171 (51) |  | 79 (47) | 53 (57) | 39 (51) | 171 (51) |  |
|  | Neutral | 7 (4) | 16 (12) | 7 (25) | 30 (9) |  | 15 (9) | 11 (12) | 4 (5) | 30 (9) |  |
|  | Agree | 25 (14) | 11 (8) | 4 (14) | 40 (12) |  | 17 (10) | 9 (10) | 14 (18) | 40 (12) |  |
|  | Strongly Agree | 2 (1) | 3 (2) | 1 (4) | 6 (2) |  | 2 (1) | 3 (3) | 1 (1) | 6 (2) |  |
|  | Strongly Disagree | 36 (20) | 26 (20) | 3 (11) | 65 (19) | 0.62 | 33 (20) | 17 (18) | 15 (20) | 65 (19) | 0.32 |
| Providing transport is my responsibility so I don't have much choice | Disagree | 74 (42) | 51 (38) | 13 (46) | 138 (41) |  | 64 (38) | 41 (44) | 33 (43) | 138 (41) |  |
|  | Neutral | 18 (10) | 23 (17) | 6 (21) | 47 (14) |  | 22 (13) | 18 (19) | 7 (9) | 47 (14) |  |
|  | Agree | 43 (24) | 30 (23) | 5 (18) | 78 (23) |  | 45 (27) | 16 (17) | 17 (22) | 78 (23) |  |
|  | Strongly Agree | 5 (3) | 3 (2) | 1 (4) | 9 (3) |  | 4 (2) | 1 (1) | 4 (5) | 9 (3) |  |
| Providing transport means I had to give up or reduced paid employment | Strongly Disagree | 70 (40) | 59 (44) | 5 (18) | 134 (40) | 0.30 | 76 (45) | 29 (31) | 29 (38) | 134 (40) | 0.07 |
|  | Disagree | 85 (48) | 57 (43) | 17 (61) | 159 (47) |  | 71 (42) | 53 (57) | 35 (46) | 159 (47) |  |
|  | Neutral | 13 (7) | 13 (10) | 5 (18) | 31 (9) |  | 18 (11) | 7 (8) | 6 (8) | 31 (9) |  |
|  | Agree | 6 (3) | 3 (2) | 1 (4) | 10 (3) |  | 3 (2) | 2 (2) | 5 (7) | 10 (3) |  |
|  | Strongly Agree | 2 (1) | 1 (1) | 0 (0) | 3 (1) |  | 0 (0) | 2 (2) | 1 (1) | 3 (1) |  |

\* Presented as 2 decimal places unless significant (<0.05).

Despite these opinions, the calls for more support and action from health professionals expressed in the focus groups were still reflected in the survey respondents as the majority felt that older drivers, especially those living in urban or rural areas, would most likely listen to health professionals about their ongoing participation in driving (p=0.0057).

## DISCUSSION

Using focus groups and community-wide surveys, this study is the first to describe the opinions older drivers, family members and caregivers have towards the NSW older driver licensing system and how they navigate it after the 2008 reforms. When viewed through the lens of the Integrative Public-Policy Acceptance framework, most participants in the focus groups and surveys were accepting of the system and held positive views towards the medical and on-road driving assessments, which they regarded as suitable and fair ways of keeping older drivers and other people safe on the roads. However, to ensure that acceptance is maintained, communication about how modified licences are set, what the medical and driving assessments are testing and how and why they were created could be improved. These sentiments have been reflected in previous studies focused on exploring the experiences of older adults who have stopped driving. Such studies have stressed the importance of increasing support and communication about the licensing regulations and alternative transport options to help older adults and their carers more effectively plan for life after.(9–11)

Licence renewal is a stressful period for both the older driver and their families and caregivers. A plethora of studies have explored how inflexible and stringent licence renewal policies have contributed to premature and ill-planned driving cessation in older drivers, especially amongst older women and those with health conditions.(12–16) Providing older drivers with clear information about what to expect may help combat stress and anxiety. This was a key message of the first theme from the FGDs with older drivers and was supported by 1/3 of the survey respondents. As mentioned in the focus groups, the on-road driving assessment is the component most are wary of, and think is the most important step in the renewal procedures. Communication about what the assessment is examining may reduce any anxiety older drivers feel, especially for those who are either older, identify as a woman, have never undertaken a driving test, have lower self-reported driving confidence or greater self-imposed driving restrictions.(17, 18) Even though the NSW licencing authority already does send out letters to older drivers about their upcoming driving assessments, not all individuals are receiving this letter as evident from the surveys. It could be argued that private driving instructors can provide information about the assessment. However, not all older drivers can afford this service, which further highlights how important it is for transport and licencing authorities to provide free and widely accessible information.

It is clear from the surveys and focus groups that family and caregivers play a vital role in helping older drivers navigate the licensing system, plan for driving cessation, and remain mobile after driving retirement; opinions which have also been expressed in previous studies looking at how older adults transition to driving retirement.(19–22)

However, support given to families and caregivers to assist older drivers is lacking. Caregivers in a recent Canadian study felt that too much burden was placed on them to provide transport for non-driving older adults and that there was little to no support from the licensing and public transport authorities.(10) This same sentiment was expressed in this study where approximately 60% of the surveyed family members and caregivers felt that older adults would listen more to the advice of a health professional and person of authority and called for a more holistic approach to older driver licensing.

This idea of a shared responsibility has been brought up before by older drivers themselves who expressed that individual, interpersonal, societal and government systems should work together to support older adults who have lost their licence.(23) This approach would best work if it was individually tailored which connects to the idea that all individuals age differently and have different needs.

The call for transparency in how evidence is presented, and regulations are created was also stressed by the family and caregivers in the focus group. However, care should be taken about the level of detail that is released to the public. Research has shown that increasing the level of detail provided about a policy may make someone understand the policy less. Instead, providing less detailed information in a segmented format where content is categorised into key points may be the best way to increase policy understanding.(24) There is therefore scope for future work to look at how best to disseminate future resources and timely communication to the public.

There was confusion around how modified licences are obtained and how their distance restrictions are set. This is of concern as by the time the groups were being conducted, eight years had passed since modified licences were introduced.

Restricting one’s driving distance is one the most common ways people modify and self-regulate their driving to gradually plan for driving retirement and healthy ageing.(25)

For older drivers with age-related medical conditions, driving restrictions, which include decreasing driving mileage and radius, have been shown to decrease crash risk, reduce traffic violations and potentially increase driver mobility.(26) Modified licence holders in NSW are approximately 20% less likely to be involved in a crash than those with full, unrestricted licences.(6) It is therefore vital for older drivers and their families and carers to know and understand more about modified licences and how they can help with a smoother transition to driving retirement.

There differences in viewpoints for the perceived benefits of the medical and driving assessments for themselves versus others. For themselves, the assessments were a way to self-check their driving abilities as they could adequately detect changes in their own driving skills (Theme 1). Since others were not as capable of gauging their driving abilities, the assessments are instead tools to identify and “catch” those who should not be behind the wheel (Theme 2). This is known as the third person effect leading towards paternalistic policy support.(27) In the context of road safety, the support for the older driver licencing policy is driven by the belief that it will benefit others who are more susceptible to unsafe driving behaviours and habits rather than themselves. This may be because older drivers tend to rate their driving abilities as better than their younger self, same-aged peers and younger drivers.(28) However, family members tend to be more reserved with their ratings. They believe all older drivers experience age-related declines in function which make them less safe behind the wheel, especially as modern traffic conditions become more complex.(29) This might explain why family members and caregivers in the survey believed older drivers would be more nervous about the assessments than the older drivers themselves. Discrepancies between the older drivers and their family and friends on how they view the assessments can impact how conversations on planning for driving retirement are approached. Since supportive statements and reappraisals from individual family members have been identified as the most effective ways to approach the topic of driving cessation,(22, 30) future research can focus on incorporating these strategies into resources and programs to help families more effectively communicate with and support both driving and non-driving older adults.

Strengths of this study come from its use of a mixed-method approach to explore concepts in depth but also systematically document the prevalence of different opinions of older drivers and family members and caregivers. By using the themes from the focus groups to develop the survey, it allowed the questions to be person-centred and reflective of real-world perspectives of people navigating aged-based licensing.

Gaps identified in the focus group, such as misunderstandings on modified licences and the driving assessment, were prioritised and expanded upon in the survey questions. The conduct of the focus groups in both metropolitan and regional areas also meant the findings could be generalised to a greater population. Similarly, the community-wide reach of the survey and the quotas on age, gender and urbanisation meant that the surveys’ results were more reflective of the population of NSW at that time. However, there are limitations to acknowledge. The first is that more focus groups were conducted in urban than regional centres which may impact the relevance of the findings to drivers and family members and caregivers outside of metropolitan Sydney. However, the authors believe the effects of this may have been lessened as the surveys included quotas for those in rural and remote regions. Survey responses also relied on self-reports which are subject to recall bias and may not accurately reflect how the respondent was thinking as they went through licensing system. The survey did not consider how long it had been since respondents went through the licensing renewal system which could have impacted how they answered the questions and affected the data’s validity. Self-selection bias may also impact the results as there is a possibility that only those who had more overwhelmingly positive or negative experiences with the licensing system were willing to participate. There were also no questions in both the FGD guides and the surveys which explored how costs associated with licensing renewals may impact how the aged-based licensing system is received.

In conclusion, this study shows that older drivers and their families and caregivers are accepting but have constructive suggestions about implementation of age-based licensing in NSW, after the 2008 reforms. Confusion about the modified licences and what the medical and driving assessments are testing highlight the need for ongoing support and engagement with the whole licensing system and pathways to driving retirement. Families and caregivers wanted more transparency on the evidence and voiced their preference for a support system with more engagement and support from licensing authorities and health professionals. The findings from this study may help develop communication channels aimed at making the age-based licensing system an easier system to navigate for all older drivers and their families and caregivers.

## Data Availability

All data produced in the present study are available upon request to the authors.

## Acknowledgements

The authors would like to acknowledge A/Prof Sheree Bekker and Dr Aryati Yashadhana for their work and assistance on coding the transcripts. We would like to further thank Transport for New South Wales for this support in this work.

## Notes

### Competing Interest Statement

Financial support was provided by the Centre for Road Safety, Transport for New South Wales, Australia to Lisa Keay. All other authors declare no competing financial or personal interests.

### Funding Statement

This work was supported by the Centre for Road Safety, Transport for New South Wales, Australia.

### Author Declarations

Human research ethics committee of the University of Sydney gave ethical approval for this work.

