## Supplemental Table 1 for "Older drivers, family members and caregivers’ views on the NSW age-based licensing; a mixed-methods approach"

| **Characteristic** | **N** | **n (%)** |
| --- | --- | --- |
| Age group, years | 608 |  |
| 65-74 |  | 209 (34.4) |
| 75-84 |  | 199 (32.7) |
| 85+ |  | 200 (32.9) |
| Sex | 608 |  |
| Male |  | 291 (47.9) |
| Female |  | 317 (52.1) |
| Region | 608 |  |
| Urban |  | 303 (49.8) |
| Rural |  | 203 (33.4) |
| Remote |  | 102 (16.8) |
| Licence type | 558 |  |
| Car |  | 499 (89.4) |
| Rider |  | 21 (3.8) |
| Rigid/Combination |  | 68 (12.2) |
| Licence status | 608 |  |
| Full |  | 537 (88.3) |
| Modified |  | 21 (3.5) |
| Retired |  | 50 (8.2) |
| Licence conditions | 185 |  |
| Glasses |  | 169 (91.4) |
| Distance restricted |  | 21 (11.2) |
| Other |  | 8 (4.3) |
| Medical Condition |  | 22 (11.9) |
| Alternate transport use | 608 |  |
| Several times/week |  | 45 (7.4) |
| At least once/week |  | 51 (8.4) |
| At least fortnightly |  | 30 (4.9) |
| Once/month |  | 47 (7.7) |
| Once/2 months |  | 24 (3.9) |
| Less often |  | 150 (24.7) |
| Never |  | 261 (42.9) |
| Driving space | 558 |  |
| Immediate neighbourhood |  | 514 (92.1) |
| Beyond neighbourhood |  | 345 (61.8) |
| More distant towns/suburbs |  | 229 (41.0) |

**Suppl. Table 1** Characteristics of older drivers surveyed in the NSW community telephone survey
