## Supplemental Table 2 for "Older drivers, family members and caregivers’ views on the NSW age-based licensing; a mixed-methods approach"

| **Characteristic** | **N** | **n (%)** |
| --- | --- | --- |
| Age group, years | 601 |  |
| 45-54 |  | 104 (17.3) |
| 55-64 |  | 180 (30.0) |
| 65+ |  | 317 (52.7) |
| Sex | 602 |  |
| Male |  | 224 (37.2) |
| Female |  | 378 (62.8) |
| Region | 602 |  |
| Urban |  | 298 (49.5) |
| Rural |  | 240 (39.9) |
| Remote |  | 64 (10.6) |
| Older driver age, years | 602 |  |
| 65-74 |  | 240 (39.9) |
| 75-84 |  | 201 (33.4) |
| 85+ |  | 161 (26.7) |
| Older driver sex | 602 |  |
| Male |  | 331 (55.0) |
| Female |  | 271 (45.0) |
| Older driver licence type* | 559 |  |
| Car |  | 528 (94.4) |
| Rider |  | 21 (3.8) |
| Rigid/Combination |  | 58 (10.4) |
| Not sure |  | 3 (0.5) |

**Suppl. Table 2** Characteristics of family members and caregivers surveyed in the NSW community telephone survey
